# Mapping insecticide resistance in mosquitoes to aid malaria control

**DOI:** 10.1101/2020.04.01.20049593

**Authors:** Catherine L. Moyes, Duncan Kobia Athinya, Tara Seethaler, Katherine Battle, Marianne Sinka, Melinda P. Hadi, Janet Hemingway, Michael Coleman, Penelope A. Hancock

**Affiliations:** Big Data Institute, Li Ka Shing Centre for Health Information and Discovery, University of Oxford, Oxford OX3 7LF, UK; Vestergaard Frandsen (EA) Ltd, Nairobi, Kenya; Clinton Health Access Initiative, Boston, Massachussetts, United States of America; Department of Zoology, University of Oxford, Oxford OX1 3RB, UK; Vestergaard SA, Lausanne, Switzerland; Department of Vector Biology, Liverpool School of Tropical Medicine, Liverpool L3 5QA, UK

**Keywords:** pyrethroid, long-lasting insecticide-treated nets, indoor residual spraying, insecticide resistance management, metabolic resistance

## Abstract

Malaria vector control may be compromised by resistance to insecticides in vector populations. Actions to mitigate against resistance rely on surveillance using standard susceptibility tests, but there are large gaps in the monitoring data. Using a published geostatistical ensemble model, we have generated maps that bridge these gaps and consider the likelihood that resistance exceeds recommended thresholds. Our results show that this model provides more accurate next-year predictions than two simpler approaches. We have used the model to generate district-level maps for the probability that pyrethroid resistance in *Anopheles gambiae* s.l. exceeds the World Health Organization (WHO) thresholds for susceptibility and confirmed resistance. In addition, we have mapped the three criteria for the deployment of piperonyl butoxide-treated nets that mitigate against the effects of metabolic resistance to pyrethroids. This includes a critical review of the evidence for presence of cytochrome P450-mediated metabolic resistance mechanisms across Africa. The maps for pyrethroid resistance are available on the IR Mapper website where they can be viewed alongside the latest survey data.

**Significance Statement:** Malaria control in Africa largely relies on the use of insecticides to prevent mosquitoes from transmitting the malaria parasite to humans, however, these mosquitoes have evolved resistance to these insecticides. To manage this threat to malaria control, it is vital that we map locations where the prevalence of resistance exceeds thresholds defined by insecticide resistance management plans. A geospatial model and data from Africa are used to predict locations where thresholds of resistance linked to specific recommended actions are exceeded. This model is shown to provide more accurate next-year predictions than two simpler approaches. The model is used to generate maps that aid insecticide resistance management planning and that allow targeted deployment of interventions that counter specific mechanisms of resistance.

## Introduction

Malaria control is primarily achieved by targeting the mosquito vectors that transmit the malaria parasite. In Sub-Saharan Africa, vector control relies heavily on insecticides integrated into bednets or applied to indoor walls (1). This means that vector control, and thus malaria control, may be compromised by resistance to these insecticides in vector populations (2).

The World Health Organization (WHO) published a Global Plan for Insecticide Resistance Management in Malaria Vectors (hereafter the “Global Plan for IRM”) that outlines best practice strategies for preserving or prolonging the effectiveness of the insecticides used for malaria control, based on the best available evidence at the time (3). Recommended actions depend on surveillance of insecticide resistance measured by standard insecticide susceptibility tests as summarised in Table 1. These actions rely on the availability of insecticides with distinct modes of action, that can be either rotated annually, applied in spatial mosaics, used in combinations of different interventions, or applied as mixed formulations. For indoor residual spraying, four neurotoxic insecticide classes have been used over many decades (carbamates, organochlorines, organophosphates and pyrethroids) and, more recently, a neonicotinoid has also been approved (4). In contrast, the range of insecticides recommended for insecticide-treated nets is more restricted, and until 2016 was limited to pyrethroids, either alone or in combination with piperonyl butoxide (PBO), a synergist that counters P450 cytochrome-mediated resistance to pyrethroids, but does not have an impact in the absence of this mechanism of resistance.

**Table 1.** Recommendations regarding insecticide choice for long-lasting insecticide-treated bednets (LLINs) and indoor residual spraying (IRS).

| 1. What is the primary intervention currently in use? | 2: What is the resistance status (susceptibility test result)? |  |  |  |
| --- | --- | --- | --- | --- |
| | Susceptible<br>($>98\%$ mortality) | Pyrethroid resistant<br>($<90\%$ mortality) | | |
| | | No cytochrome P450-mediated resistance or no data<br>($<10\%$ increase in mortality with PBO) | Cytochrome P450-mediated resistance<br>($\geq 10\%$ increase in mortality with PBO) | |
|  |  |  | (10-80% mortality without PBO) | (mortality without PBO not 10-80%) |
| **LLINs** | Continue monitoring using susceptibility tests and, if cases are increasing, ensure timely replacement of worn-out nets and assure the quality and extent of LLIN coverage. | In addition to LLINs, introduce focal IRS in annual rotations, at least in areas of greatest concern, and use non-pyrethroid active ingredients when they become available, providing this does not compromise coverage. | Consider switching to PBO-treated LLINs. and/or introducing focal IRS in annual rotations, at least in areas of greatest concern, and using non-pyrethroid active ingredients, providing this does not compromise coverage. | Introduce focal IRS in annual rotations, at least in areas of greatest concern, and use non-pyrethroid active ingredients, providing this does not compromise coverage. |
| **IRS** | Pre-emptive rotation of insecticide classes with confirmed susceptibility. | Switch away from current insecticide and rotate alternative compounds. |  |  |

This is an abridged summary of the recommendations in Tables 4 and 5 of the Global Plan for IRM and accompanying text, the thresholds defined in the WHO's Test Procedures for Insecticide Resistance

Long-lasting insecticide-treated nets (LLINs) are the primary recommended intervention for vector control and the use of pyrethroid-treated nets has underpinned the reductions in malaria prevalence from 2000 to 2015 (1, 5). For this reason, the Global Plan for IRM places particular emphasis on prolonging the effectiveness of pyrethroids in vector control. All currently recommended insecticide-treated net formulations use pyrethroids, but dramatic increases in pyrethroid resistance in the major African malaria vectors have occurred from 2005 to 2017 (4, 6). Fortunately, three partner compounds have recently been introduced to net formulations. WHO prequalification (PQ) status has been awarded to LLINs impregnated with a combination of the pyrethroid, α-cypermethrin, and the pyrrole, chlorfenapyr (7), and to LLINs impregnated with α-cypermethrin and the insect growth regulator, pyriproxyfen (8). Long-lasting insecticide-treated nets treated with a pyrethroid (a- cypermethrin, deltamethrin or permethrin) in combination with PBO first became available ten years ago and in 2017 the WHO gave pyrethroid-PBO nets an interim endorsement as a new vector control class pending further review of the epidemiological data from randomised control trials (9). The WHO’s Global Plan for IRM covers the use of alternative insecticide classes, but does not provide guidelines for use of products that contain a synergist that counters a specific mechanism of resistance. The WHO, therefore, published the Conditions for Deployment of Mosquito Nets Treated with a Pyrethroid and Piperonyl Butoxide (hereafter the “Conditions for Deployment of PBO-treated Nets”) in 2017 (10, 11). These guidelines also recommend actions that depend on surveillance of insecticide resistance measured by standard insecticide susceptibility tests (Table 1).

Monitoring, and the recommendations outlined in the Conditions for Deployment of PBO-treated Nets (3, 10, 12). The recommendations linked to the presence of *kdr* mechanisms have been excluded for reasons of brevity and because these are not materially different from ‘resistance without mechanism data’ or ‘no cytochrome P450-mediated resistance’. The threshold values for susceptibility test results are given in parentheses.

Although the current recommendations in the Global Plan for IRM and the Conditions for Deployment of PBO-treated Nets rely on surveillance of insecticide resistance measured by standard insecticide susceptibility tests (12, 13), a 2017 review of implementation of the Global Plan for IRM found monitoring was inadequate and inconsistent in most settings (14, 15). A review of the data available today shows that this is still true (Figure 1).

**Figure 1.**
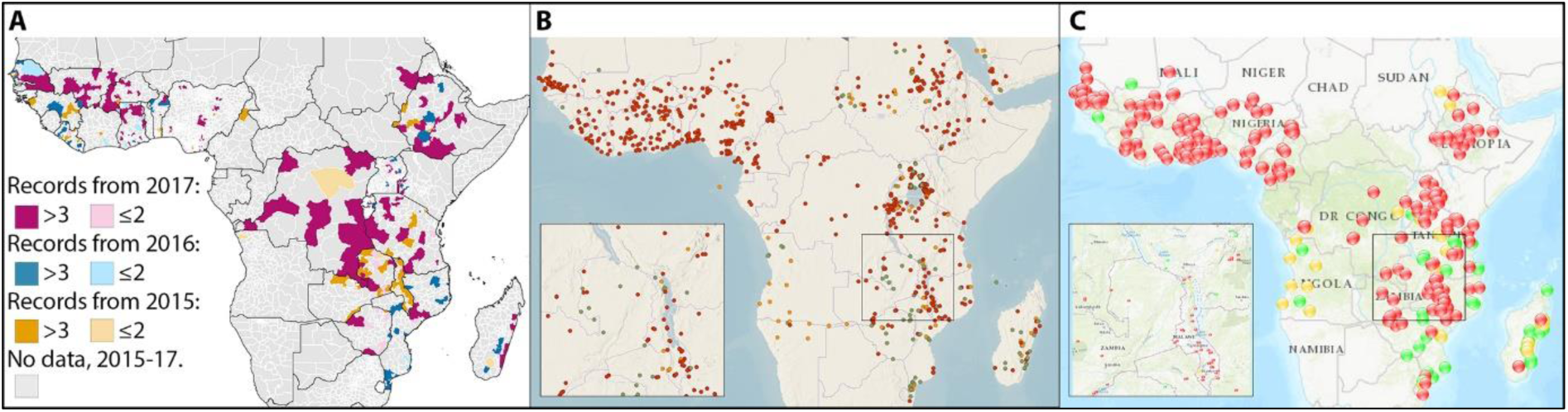
Gaps in the available insecticide resistance surveillance data. (**A**) Districts with susceptibility test (bioassay) results for 2015-2017 from a published susceptibility test dataset that collated data from routine monitoring by agencies such as the President’s Malaria Initiative (PMI) and from research groups in Africa, and was subject to rigorous quality assurance processes (15). Ninety four percent (3,136/3,323) of malaria- endemic second order administrative divisions do not have any data for insecticide resistance in this year and 89% (2,962/3,323) do not have any data in this year or the two preceding years. (**B**) Data shown on the WHO Malaria Threats website for the years 2015-2019 (accessed 9 December 2019) (16). (**C**) Data shown on the IR Mapper website for the years 2015-2019 (accessed 9 December 2019) obtained from published articles and PMI surveillance (17).

To address these issues, we propose using the results of a recently developed geostatistical ensemble model that was fitted to a published series of susceptibility test surveillance data covering 31 African countries and spanning a period of 13 years (6, 15). The geostatistical ensemble model accounts for noise in the susceptibility test data and provides a set of thirteen fine-resolution, annual maps for each year from 2005 to 2017, representing the mean prevalence of resistance in *Anopheles gambiae* s.l. to each of the pyrethroids most commonly used in LLINs (α-cypermethrin, deltamethrin, permethrin, and λ-cyhalothrin) and resistance to DDT. These maps provide values in data sparse areas and are potentially a powerful tool for use when considering the choice of insecticide for malaria vector control. In addition to gaps in the surveillance data, decision-makers are often faced with data that are not up-to-date (Figure 1). Here we demonstrate the capacity of the geostatistical ensemble model to predict pyrethroid resistance one year ahead, out-performing simple district-level data summaries. Tools for decision makers need to incorporate threshold values that are linked to specific actions (Table 1) and provide information about the uncertainty around the predictions for achieving these thresholds. We demonstrate how the published geostatistical ensemble model can be used to calculate the probability of these thresholds being met. In addition to the key insecticide susceptibility thresholds defined by resistance management plans, the deployment of PBO-treated nets requires evidence for the presence cytochrome P450-mediated mechanisms of resistance (Table 1) (10). Here we therefore also review and map the evidence for presence of these mechanisms of resistance. Finally, we present a set of maps that show the probability of exceeding thresholds linked to recommended actions at a district scale, i.e. the spatial resolution commonly used for operational decisions.

## Material and Methods

### Identifying gaps in the surveillance data

Standard susceptibility test data for pyrethroid resistance in all African malaria vectors were extracted from the largest publicly-available database of insecticide resistance data (15) and processed in QGIS 3.4.12 software. Each data point was linked to a year and to either the geographical coordinates for a precise collection location or an identifier for the administrative unit where the mosquitoes were collected. These two pieces of information were used to calculate the number of data points in each year in each second order administrative unit (hereafter referred to as districts). Each district was also classified as malaria-endemic or not by overlaying the administrative unit layer with the raster maps of *Plasmodium falciparum* and *P. vivax* that are available from the Malaria Atlas Project website (18). Non-endemic areas were discarded and the proportion of endemic districts with susceptibility test data for each year was calculated. The resulting map is presented alongside the most recent data from the IR Mapper and Malaria Threats websites (16, 17).

### Existing geostatistical ensemble model used in this study

This study used the outputs from a published Bayesian geostatistical ensemble model (6), which were used here to generate the cumulative probability maps described below. Further, the model was re-run to test its ability to provide next-year predictions. The ensemble model has been described in full by Hancock *et al*. (6). Briefly, the Bayesian geostatistical ensemble model was fited to over 6,000 data points from susceptibility tests that exposed *An. gambiae* s.l. mosquitoes to four pyrethroids and DDT. The model leveraged (i) associations between resistance and potential explanatory variables such as LLIN coverage, and (ii) spatial patterns in the resistance data. This ensemble model generated a posterior distribution of predictions for every cell in a fine resolution (2.5 arc minute, approximately 5km) grid. Validation of the outputs by Hancock *et al*. using withheld data (out-of-sample validation) showed that the ensemble model produced robust predictions within the area and time-frame for which data on pyrethroid resistance were available (6). For the current study we used the model output for mortality after deltamethrin exposure in a standard susceptibility test. We selected deltamethrin resistance because this pyrethroid has the largest volume of susceptibility test data available, and the deltamethrin resistance outputs had the narrowest credible intervals (6). There are, however, strong associations among the patterns of resistance to all four pyrethroids (α-cypermethrin, deltamethrin, permethrin, and λ-cyhalothrin) (19).

### Calculating district-level resistance means

To generate maps of mean mortality across each district 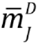, we used the mean values produced by the model for every grid cell, weighted using values for *An. gambiae* s.l. habitat suitability that are available at the same resolution ∼5km as the resistance values (20):

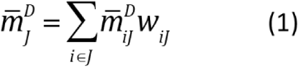

Where 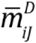 is the predicted mean mortality for deltamethrin in grid cell *i* and *w*_*iJ*_ is relative habitat suitability.

### Calculating the probability of a district exceeding a set mortality threshold

We created a series of maps for the probability of meeting the resistance thresholds used in management decision-making (Table 1), at the district level. We used the full posterior distributions of resistance predictions made by the geostatistical ensemble model to produce maps of (i) the probability that the mean mortality was ≥98% (the WHO threshold for “susceptibility) and (ii) the probability that the mean mortality was <90% (the WHO threshold for “resistance”).

The posterior distribution of the weighted district mean (equation 2) was calculated using *k*=1000 draws from the posterior distribution of the predicted resistance in each pixel, 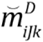, and then calculating a weighted mean, 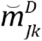, across each district for each posterior draw:

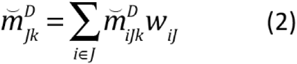

The cumulative probabilities (i) and (ii) across all posterior draws were then calculated. A wider range of thresholds is useful to visualise the areas worst affected by resistance so we also calculated the cumulative probabilities that each district has a mean mortality of <80%, <70%, <60%, <50%, <40%, <30%, <20% and <10%.

### Assessing whether the criteria for deployment of piperonyl butoxide-treated nets have been met

Deployment of pyrethroid LLINs treated with piperonyl butoxide (PBO) is currently recommended in places that meet all of the following criteria: i) there is pyrethroid resistance that; ii) results in 10- 80% mortality in a susceptibility test; and iii) is conferred (at least in part) by a monooxygenase-based resistance mechanism (that is, elevated levels of cytochrome P450) (10). To test which districts meet the first two criteria, we used the full posterior distributions of predictions to calculate the probability of the mean mortality being 10-80% for each district, as described above for the 98% and 90% threshold maps.

To assess the third criterion, results of paired standard pyrethroid susceptibility tests conducted from 2005 to 2018 with and without PBO were extracted from a published database (15). The WHO protocol for these tests defines a difference of ≥10% as evidence for the presence of cytochrome P450-mediated mechanisms of resistance (12). The mortality obtained with the addition of the synergist was plotted against mortality after exposure to the pyrethroid alone, and the data were divided into those pairs with a difference of ≥10% mortality and those with a difference of <10% mortality. Each data point had a link to geographical coordinates for the location of mosquito collection and these were used to associate each data point with a second order administrative unit using the QGIS 3.4.12 software. We used these data to generate a map that classified each district tested according to whether it had evidence for presence of this mechanism, or for its absence, or no evidence. In some instances, more than one data point was linked to a single district. If a single district had a test result(s) that showed the presence of this mechanism, as well as a test result(s) that showed an absence of this mechanism, then the district was classified as having evidence for the presence of cytochrome P450-mediated mechanisms of resistance.

### Assessing whether predictive maps can compensate for a lack of contemporary data

One limitation of the available information on resistance, whether it consists of susceptibility test data or modelled predictions for mean test results, is the time lag between mosquito specimens being collected in the field and tested for resistance, and the data being checked, compiled and made available. The most up-to-date information available is inevitably out-of-date once it has been cleaned and collated (Figure 1), but decision-makers require information on the current situation. We compared our pyrethroid resistance maps to the observed mortality values one year ahead. We also compared the values obtained using two simpler methods to the observed mortality values one year ahead. The two simpler methods involved calculating (i) the mean, and (ii) the minimum value of the susceptibility test results from the current year for each district. We assessed accuracy using next-year out-of-sample validation. For example, we used the geostatistical ensemble modelling approach described in Hancock et al. (2019) to predict resistance based on a subset of the susceptibility test data spanning the time period 2010-2014, and compared the predictions for 2014 to the withheld test data available for 2015. Similarly, values for 2015 and 2016 were generated using models fitted to data subsets spanning the time periods 2010-2015 and 2010-2016, then compared to the observed data for 2016 and 2017, respectively. For values calculated using simpler district-level data summaries, we produced summaries of the susceptibility test data available for each district for the current year (2014, 2015 and 2016 individually), and then compared these to the next-year observed values for 2015, 2016 and 2017 respectively. We compared the root mean square error (RMSE) across each of the three methods, for each of the three years predicted, for each geographical region (west and east Africa) to give 6 comparisons of predictive performance.

The geostatistical ensemble model also has the advantage of being able to provide modelled resistance values for districts in which no susceptibility test data for the current year are available. We calculated additional RMSE values comparing the modelled values for districts lacking current-year data with the next-year observed data for the same three pairs of consecutive years described above.

### *Overlaying the geographical distributions of* An. gambiae *s.l. and* An. funestus

The model described above predicted resistance in *An. gambiae* s.l. and this complex includes three widespread and important malaria vector species (*An. arabiensis, An. coluzzii* and *An. gambiae*), however, *An. funestus* is also amongst the most important malaria vector species in Africa. When used in decision-making, the model results generated by this study need to be considered alongside the respective distributions of the *An. gambiae* complex and *An. funestus*. To produce a single map of their respective distributions, predictive maps of the relative probability of occurrence of *An. gambiae* s.l. and *An. funestus* produced by Wiebe *et al*. (2016) were converted to binary maps of presence and absence. The threshold predicted value from the *An. gambiae* s.l. map that captured 95% of the confirmed *An. gambiae* s.l. occurrences was identified and used to convert the continuous value maps into binary versions. The value of 95% of confirmed reports was selected to allow for errors in the confirmed reports, including errors in geopositioning of sites and errors in species identification. This process was repeated to generate a binary *An. funestus* map. The two binary data layers were then overlaid to provide a single map showing where both *An. gambiae* s.l. and *An. funestus* are likely to occur.

## Results

### Mapping the probability of districts exceeding a set mortality threshold

We provide a set of three district-level maps to assist decision-makers in assessing pyrethroid resistance levels and evaluating each district with respect to the resistance thresholds stated in policy guidelines (Figure 2). The three maps are the weighted mean resistance to deltamethrin for each district (Figure 2a), the probability of mortality to deltamethrin being less that the WHO threshold for susceptibility (Figure 2b), and the probability of mortality to deltamethrin exceeding the WHO threshold for confirmed resistance (Figure 2c). The latter two maps aim to communicate the uncertainty associated with evaluating whether each district meets these thresholds, where the highest and lowest probability values indicate certainty whereas intermediate probabilities indicate uncertainty (Figures 2c and 2d). The results show that the threshold for susceptibility is highly unlikely to be met in any district in west or northeast Africa in 2017, but some districts in southern east Africa had probabilities of greater than 0.5 for meeting the susceptibility threshold (Figure 2b). The threshold for confirmed resistance was highly likely to be met in most districts in west Africa in 2017, although some districts had an intermediate probability (Figure 2c). In southern east Africa there was an intermediate or low probability of confirmed resistance. These maps show resistance in the *An. gambiae* species complex and the distribution of this complex is shown in Figure 2d alongside the predicted distribution for *An. funestus*. The results shown in Figures 2a-c are relevant in all areas where *An. gambiae* s.l. is present (Figure 2d) but in areas where *An. funestus* is also present, these results should be considered alongside data on resistance in *An. funestus*. The published threshold values that are linked to defined actions may change as new evidence becomes available. In addition, different thresholds may be selected where resources for more expensive interventions are limited meaning that they can only be deployed in the worst-affected areas. For these reasons, maps for thresholds at every 10% interval were also generated and the animation in Supplementary File 1 shows the probability of the mean mortality for a district being below each threshold value from 90% to 10% mortality.

**Figure 2.**
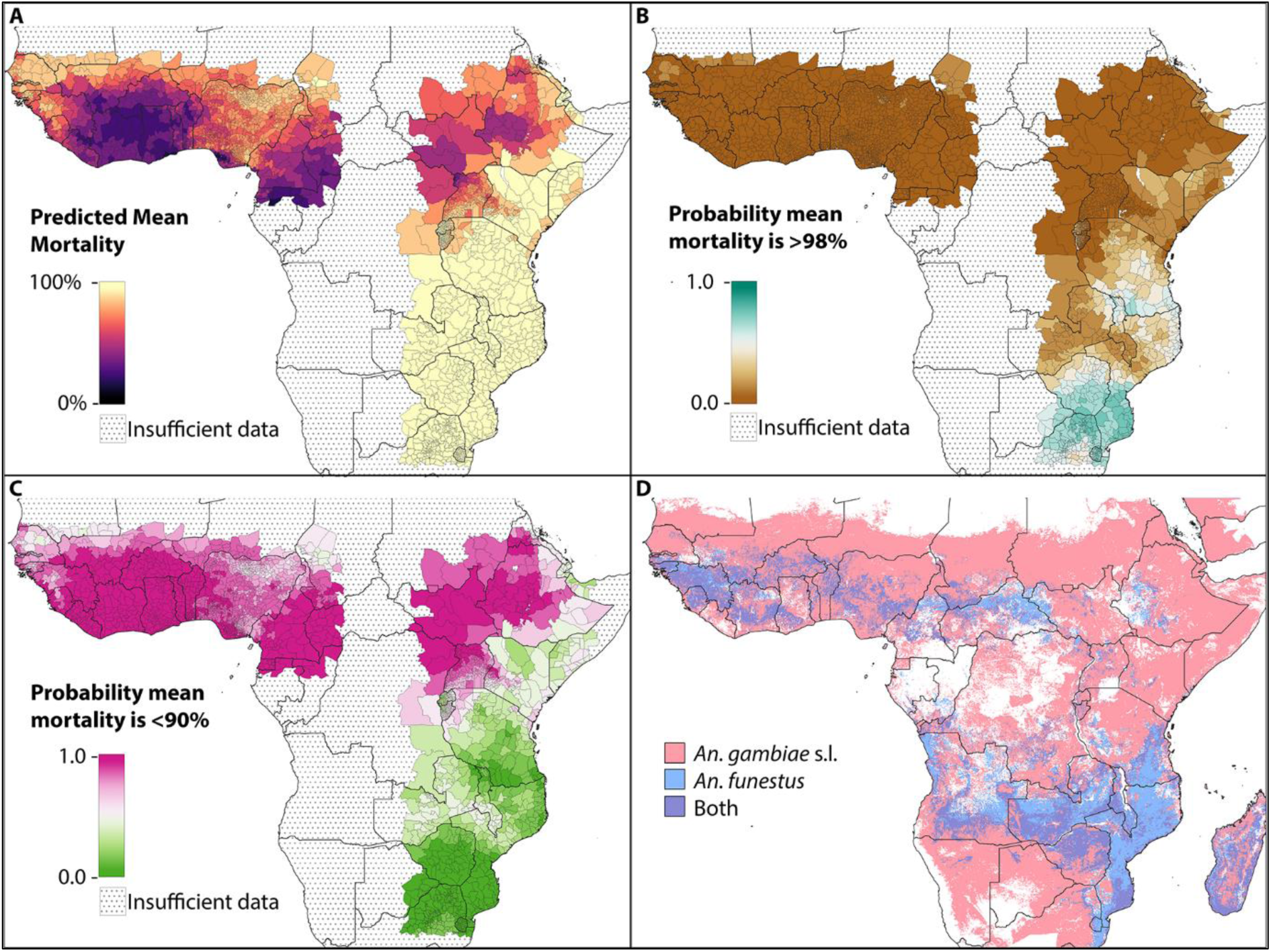
District-level deltamethrin resistance in local *An. gambiae* s.l. populations. (**A**) Predicted mean susceptibility test mortality for each district in 2017. (**B**) Probability that mean susceptibility test mortality for the district is >98%, the definition of a “susceptible” population. (**C**) Probability that mean susceptibility test mortality for the district is <90%, the definition of a “resistant” population (**D**) The presence of *An. gambiae* s.l. and/or *An. funestus* based on previously published baseline species distributions (20).

### Assessing whether the criteria for deployment of piperonyl butoxide-treated nets have been met

Deployment of pyrethroid LLINs treated with piperonyl butoxide (PBO) is currently recommended in places that meet all of the following criteria: i) there is pyrethroid resistance that; ii) results in 10- 80% mortality in a susceptibility test; and iii) is conferred (at least in part) by a monooxygenase-based resistance mechanism (this is, elevated levels of cytochrome P450), providing deployment does not compromise the levels of LLIN coverage that are achieved (10). The predicted probability that district-level mortality after deltamethrin exposure is 10-80% shows that deployment of these nets would currently be recommended in much of west and northeast Africa if cytochrome P450- mediated mechanisms of resistance were confirmed in these districts (Figure 3a). Data for the presence of these mechanisms (defined by WHO in note i of Figure 3.1 of the 2016 test procedures as a ≥ 10% increase in susceptibility test mortality after the addition of PBO (12)) show that 79% (276/351) of the surveillance data for *An. gambiae* s.l. confirms the presence of these mechanisms, as does 94% (34/36) of the data for *An. funestus*. All 20 countries that were tested had evidence of elevated cytochrome P450 and the most recent evidence within each country was always for presence. The most recent year tested for these countries ranged from 2009 to 2018. When these data are plotted geographically, no obvious geospatial trends are apparent (Figure 3b). It is also clear that there is variation in the increase achieved when PBO is used, from 10-100%, and considerable noise in the data (Figure 3c).

**Figure 3.**
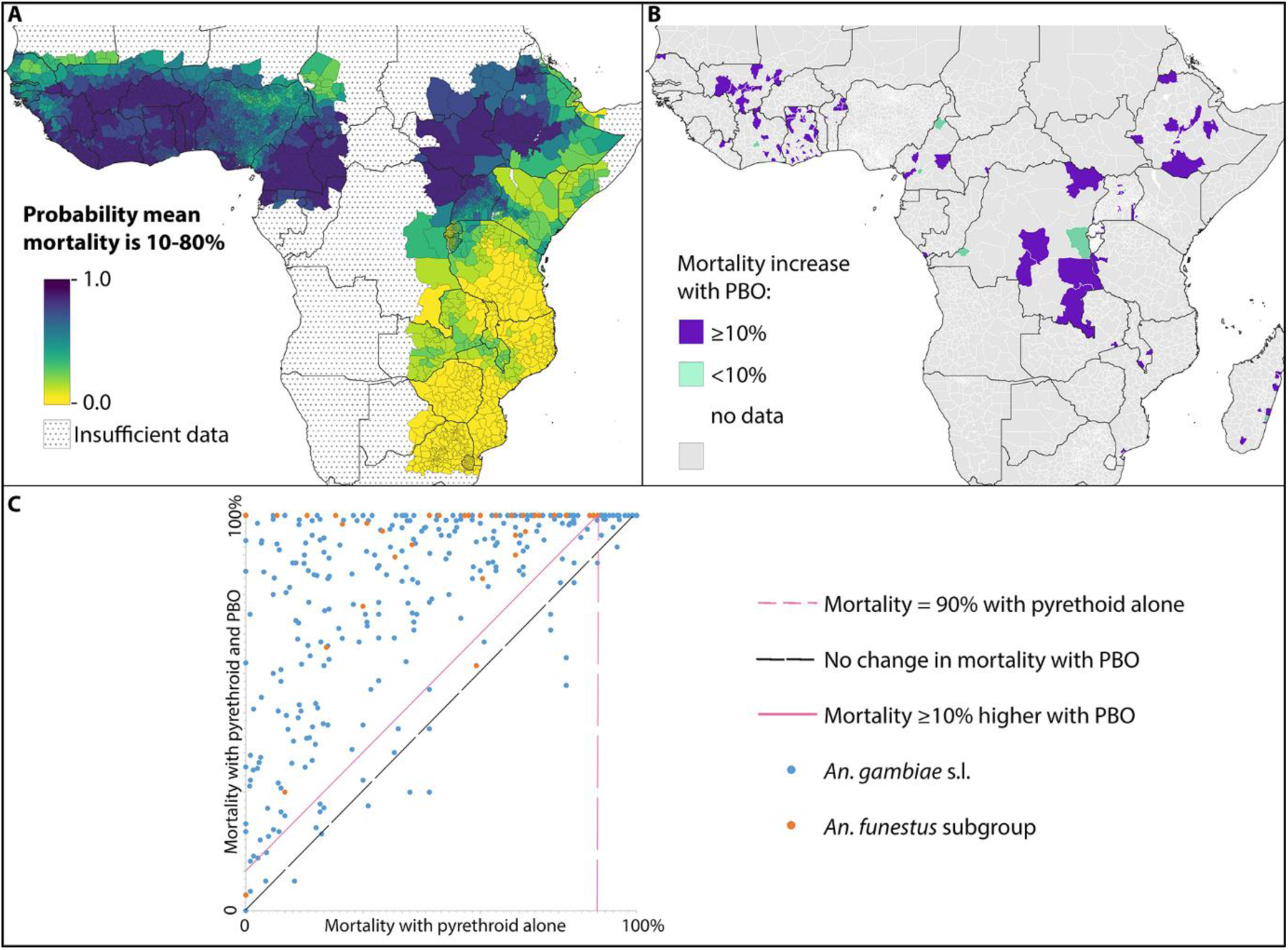
The probability of meeting the criteria for the deployment of PBO-treated nets. (**A**) The predicted probability that the mean mortality for a district is 10-80%. (**B**) Districts with surveillance data from paired standard susceptibility tests using a pyrethroid with and without the synergist PBO. (**C**) Plot of surveillance data from paired standard susceptibility tests using a pyrethroid with and without the synergist PBO showing the full variation in the results and key threshold defined in WHO guidelines.

There are substantial gaps in the susceptibility test data for cytochrome P450-mediated pyrethroid resistance with 96% (3,200/3,323) of malaria-endemic districts having no data in any year. Of the districts with data, 92% (114/123) meet the criteria for confirmed cytochrome P450-mediated pyrethroid resistance and 8% of the districts tested had no results that met these criteria. It is worth noting that 41 districts had multiple test results that gave conflicting outcomes of both greater than (presence) and less than (absence) 10% increase in mortality with the addition of PBO. That is, the majority of districts (41/50) with one or more test results that did not meet the criteria to confirm cytochrome P450-mediated pyrethroid resistance also had a test result that did meet the criteria. This may indicate variation within-district, noise in the test data, or rapid spread of this mechanism of resistance. Either way, there is clearly little definitive evidence for absence of elevated cytochrome P450 in any of the regions tested.

Collectively these results show that much of west Africa is likely to have a level of pyrethroid resistance that meets two of the deployment criteria. Most of these districts do not have cytochrome P450 mechanism data, however, the closest data point to each district generally indicates presence of this mechanism. The area of northeast Africa with a high probability of meeting the criteria for the level of pyrethroid resistance has insufficient data to comment on whether elevated cytochrome P450 occurs.

### Assessing whether predictive maps can compensate for a lack of contemporary data

The geostatistical ensemble model performed best according to the out-of-sample root mean square error (RMSE) across the three years for both the west and east regions (Table 2). Using the mean of the values from the preceding year was considerably more accurate in predicting next-year observations than using the minimum value across the district in the preceding year.

We also compared values produced by the geostatistical ensemble model for the current year, in districts that had no test results in that year, to the observed data for the next year. The out-of-sample RMSE values obtained (Table 3) overlap with those obtained across districts that did have susceptibility test data available for the preceding year (Table 3).

These results demonstrate that predictions from the geostatistical ensemble model provide more reliable up-to-date estimates of resistance compared to predictions from simple district-level data summaries, and can estimate predict mortality in districts where no recent susceptibility test data is available.

**Table 2.** Comparing the accuracy of next-year predictions when data are available for the previous year.

| Next year | Root mean square error (RMSE) |  |  |
| --- | --- | --- | --- |
|  | Geostatistical ensemble model | Mean value of surveillance data from preceding year | Minimum value of surveillance data from preceding year |
| **West region:** |  |  |  |
| 2015 | **0.224** (*n*=149) | 0.232 (*n*=149) | 0.386 (*n*=149) |
| 2016 | **0.239** (*n*=129) | 0.276 (*n*=129) | 0.309 (*n*=129) |
| 2017 | **0.302** (*n*=243) | 0.384 (*n*=243) | 0.50 (*n*=243) |
| **East region** |  |  |  |
| 2015 | **0.186** (*n*=159) | 0.194 (*n*=159) | 0.301 (*n*=159) |
| 2016 | **0.276** (*n*=85) | 0.281 (*n*=85) | 0.298 (*n*=85) |
| 2017 | **0.277** (*n*=68) | 0.323 (*n*=68) | 0.395 (*n*=68) |

The root mean square errors (RMSE) of predicted and observed next-year resistance where all data obtained after the given year were withheld from the prediction methods. A lower RMSE indicates a more accurate prediction. Observed datasets consisted of all pyrethroid susceptibility test results (performed using α-cypermethrin, deltamethrin, permethrin, and λ-cyhalothrin) from the next year for districts where test results for the preceding year were available. 'n' is the number of withheld data points. The RMSE values shown in bold indicate the best performing model for each year.

**Table 3.** Comparing the accuracy of next-year predictions when no data are available for the previous year.

| Next year | Root mean square error (RMSE) |  |
| --- | --- | --- |
|  | West region | East region |
| 2015 | 0.315 (*n*=93) | 0.218 (*n*=88) |
| 2016 | 0.327 (*n*=102) | 0.270 (*n*=70) |
| 2017 | no data | 0.289 (*n*=75) |

The root mean square errors (RMSE) of predicted and observed next-year resistance where all next-year data were withheld. A lower RMSE indicates a more accurate prediction. Observed datasets consisted of all pyrethroid susceptibility test results (performed using α-cypermethrin, deltamethrin,
permethrin, and λ-cyhalothrin) from the next year for districts where no test results for the preceding year were available. 'n' is the number of withheld data points. The West region had no districts where there was data available in 2017 but not for 2016.

### Dissemination

The stratified maps presented here and the fine resolution mean mortality maps from Hancock *et al*. (2019) (6) for the years 2005 to 2017 are available to view and download on the IR Mapper website. These maps can be viewed on the IR Mapper website alongside the susceptibility test results for the years closest to that being modelled. Users can also choose to view each of these maps overlaid with the most recent data for both *An. gambiae* s.l. and *An. funestus*, as well as other vectors. They can zoom into their geographic region of interest and export the map image they have constructed. In addition, the data files behind the stratified maps constructed during this study are available from Figshare (details to be added).

## Discussion

Here we provide maps that show the probability that the mean prevalence of pyrethroid resistance in a district meets a set of thresholds linked to specific malaria vector control recommendations. This is the first time that this information has been provided for regions with data gaps in space and time. Assessing the level of insecticide resistance is essential in order to prioritise use of products that mitigate against it (7, 8, 21, 22). The new mixed formulation malaria vector control products are more expensive to produce, and funds for malaria control are limited. Past experience with indoor residual spraying has shown that a switch to a more expensive product can reduce the coverage achieved and reductions in indoor residual spraying coverage have been linked to resurgences in malaria (23). It is therefore vital to have information such as the maps presented here that aids decisions on when and where more expensive products that mitigate against insecticide resistance are deployed (24, 25). The geostatistical ensemble model that we used has previously been shown to provide robust estimates within the area and time-frame for which sparse data on pyrethroid resistance are available (6). Here we have shown that it provides the most robust predictions for the next year, in a scenario where no data for that year are available, compared to taking the mean or minimum value of the surveillance data for the preceding year. Similar results were also obtained when no data were available for the preceding year either.

Susceptibility test data are noisy (26, 27) and the low data volumes typically seen in resistance monitoring (Figure 1) are particularly susceptible to issues of data noise. The model used here has the advantage that it separates the signal (trends in space and time) from this noise to provide mean mortality values, and this also has a smoothing effect seen in the fine resolution maps generated by Hancock *et al*. (6). The maps presented here were smoothed further by generating a set of district-wide means, and further still by setting a threshold mortality value. This results in contiguous areas that have the same resistance classification and do not always match the variation seen in the raw surveillance data, which may be the result of issues with sparse data and underlying data noise (28).

The maps presented here show the probability of resistance to deltamethrin in the *An. gambiae* species complex at a district level. The data that went into these maps were from mosquito samples representative of the *An. gambiae* species complex at each time and place, thus the maps represent resistance in the species complex at each time and place including variation caused by the combination of spatially varying species composition and differences in resistance among these species (6, 15). Other species are important, notably *An. funestus* (29-31), and use of these maps needs to be accompanied by knowledge of the locally important vectors. The maps presented here also focused on a single pyrethroid, deltamethrin. Predictions are available for α-cypermethrin, permethrin, and λ-cyhalothrin, and maps showing the probability of meeting key thresholds can also be generated for these pyrethroids. It is, however, important to note that previous work has shown strong associations among patterns of resistance to these four insecticides and evidence of resistance to one is likely to indicate resistance to the others (19). District-level maps of resistance were chosen to match the spatial resolution currently commonly used for operational decisions, but finer resolution data may increasingly be used by decision-makers in the future and the probability maps presented here can be generated down to a ∼5km resolution.

Recommendations for the deployment of PBO-treated nets are based on evidence for the mechanism of resistance as well as the level of resistance indicated by susceptibility tests (11). The most commonly available standardised data for cytochrome P450-mediated mechanisms of resistance are susceptibility tests using the PBO synergist (15, 32). More data would be required to generate predictive maps from synergist test results, however, the data review presented here poses the question of whether the benefit of having comprehensive synergist surveillance data is worth the cost of collecting such data. Here we have shown that the vast majority of locations tested are classified as having cytochrome P450-mediated pyrethroid resistance, as currently defined by WHO using synergist bioassays (12). Biochemical tests are also available but they do not perform consistently. Genetic tests give more robust results but are expensive to perform and data are therefore even more sparse. The data that are available from genetic tests do, however, show the same trends as seen for the synergist bioassay results (32).

As new evidence becomes available, many of the key definitions and thresholds that are currently recommended by the Global Plan for IRM and the Conditions for Deployment of PBO-treated Nets may be updated. The results here provide examples based on the currently recommended threshold values, however, the approach taken by our study is flexible and the supplementary files provide maps that use a full range of mortality thresholds. In addition, current policies advise that if financial constraints mean that the guidelines cannot be followed without compromising intervention coverage then the worst affected areas should be targeted. Our mean predictive value map (Figure 2a), the sliding scale of mortality thresholds (Supplementary File 1), and the continuous scale of probabilities for defined thresholds (Figure 2b, Figure 2c and Supplementary File 1) all allow users to identify such areas.

The current results are of relevance in areas where IRS is the primary intervention. In these areas, the Global Plan for IRM also recommends pre-emptive use of rotations regardless of available data on insecticide resistance but warns against making a change if it would result in reduced coverage. The cheapest insecticides are currently pyrethroids and DDT, however, cross-resistance between these two classes means that use of one can lead to resistance to the other, reducing the classes available for rotation. This means that a rotation of classes that avoids issues of pyrethroid resistance requires more expensive compounds. Despite well-reported increases in insecticide resistance, the practice of using the same insecticide class for IRS in consecutive years is still common (33). The results presented here allow groups that are deploying IRS to identify areas where pyrethroid resistance is highest to target focal use of rotations using more expensive insecticides. The same approach can be taken in areas that primarily rely on LLINs, to introduce focal IRS or LLINs treated with a non-pyrethroid active ingredient, in areas with the highest resistance.

Ultimately, decisions on which intervention to use, and which insecticide to select if applicable, depend on multiple entomological, ecological, economic, epidemiological, cultural and operational considerations (3). Future work should address this complexity using a hierarchal decision tree- based system, to simplify and standardise evidence-based decision making. Evidence of insecticide resistance and an increase in malaria cases are particularly important considerations for any such decision tree. Here we have focused on the prevalence of insecticide resistance and provide maps that can be used within a broader decision-making framework. The results generated by this study are freely available for use by all groups as stand-alone tools via the IR Mapper website or they can be obtained from Figshare for integration into more complex decision-making tools.

## Data Availability

The survey data used are available at https://www.nature.com/articles/s41597-019-0134-2.
The modelled maps are being released at http://www.irmapper.com/.

http://www.irmapper.com/

## Acknowledgements

This work was funded by Wellcome Trust Grant 108440/Z/15/Z (to C.L.M.)

## Supplementary Information

Movie file 1: This video (MP4 format) shows a consecutive series of maps for the probability that the mean mortality for each district is <10%, <20%, <30%, <40%, <50%, <60%, <70%, <80% and <90%, following *An. gambiae* s.l. exposure to deltamethrin in 2017.

